# The socioeconomic factors that led to unemployment under COVID-19 pandemic among Japanese workers: a prospective cohort study

**DOI:** 10.1101/2021.08.03.21261518

**Authors:** Makiko Kuroishi, Tomohisa Nagata, Ayako Hino, Seiichiro Tateishi, Akira Ogami, Mayumi Tsuji, Shinya Matsuda, Koji Mori, Yoshihisa Fujino, the CORoNaWork project

**Affiliations:** Department of Occupational Health Practice and Management, Institute of Industrial Ecological Sciences, University of Occupational and Environmental Health, Japan; Department of Mental Health, Institute of Industrial Ecological Sciences, University of Occupational and Environmental Health, Japan; Disaster Occupational Health Center, Institute of Industrial Ecological Sciences, University of Occupational and Environmental Health, Japan; Department of Work Systems and Health, Institute of Industrial Ecological Sciences, University of Occupational and Environmental Health, Japan; Department of Environmental Health, School of Medicine, University of Occupational and Environmental Health, Japan; Department of Public Health, School of Medicine, University of Occupational and Environmental Health, Japan; Department of Environmental Epidemiology, Institute of Industrial Ecological Sciences, University of Occupational and Environmental Health, Japan

**Keywords:** socioeconomic factors, unemployment, vulnerable workers, COVID-19, Japan

## Abstract

**Objectives:** To examine the relationship between socioeconomic factors and unemployment among workers during COVID-19 pandemic in Japan.

**Design:** A prospective cohort study, follow-up from December 22–25, 2020 through February 18–19, 2021.

**Setting:** The panelists registered with an Internet survey company

**Participants:** A cluster random sample of 33,087 workers with stratification by sex, job type, and region among Japan’s working population

**Main outcomes and measures:** Unemployment between December 2020 and February 2021

**Results:** Among the 19,941 participants, 725 (3.6%) had experienced unemployment. Multivariate analysis showed that the significant high unemployment for women (compared to men); younger age (compared to older age); being married (spouse not working), bereaved or divorced, and unmarried (compared to married (spouse working)); annual household income less than 6 million yen, (compared to more than 10 million yen); junior high or high school, vocational school, junior college, or technical school (compared to graduate school); and temporary or contract employees, self-employed, agriculture, forestry, or fishing (compared to general employees).

**Conclusions:** COVID-19 appears to have created difficulties for the vulnerable groups. This suggests the need for employment and economic support for such individuals.

**Strengths and limitations of this study:** Under the pandemic of COVID-19, we found that unemployment was associated with socioeconomic factors such as age, sex, marriage, income, education and occupation.

Our findings suggest that in the event of a major epidemic, resulting in unemployment among vulnerable segments of the labour market, regardless of whether workers themselves are infected.

It was unclear why the participants had experienced unemployment

This study was conducted as an Internet survey, so the generalizability of our results is limited.

## Introduction

Unemployment is associated with a substantial risk of poor physical and mental health. It has consistently been shown to be significantly associated with increases in chronic heart disease, acute myocardial infarction, poor mental health, mental disorders, substance-related disorders, and suicide [1-3]. Unemployment is an important factor with regard to public health: it poses a health risk for individuals; it also leads to family poverty and, ultimately, constitutes a burden on social security.

COVID-19 has spread around the world and continues to exert a profound impact on global economies. Economic activities have changed drastically as a result: people have refrained from travel and outside eating, drinking, entertainment; they are encouraged to stay at home. The resulting unemployment has become a major concern: according to the Organization for Economic Co-operation and Development (OECD), countries’ unemployment rates rose significantly around the same time as the outbreak of COVID-19 [4]. In this regard, Japan’s unemployment rate appears to have remained relatively low—around 3%. However, Official data on the unemployment rate may be underestimated, as it is defined as “people looking for work. Actually, data for Japan clearly show a marked decline in the number of people in employment and a fall in household income [5,6].

With disasters and crises in the past, the risk of unemployment was found to be particularly high among socially vulnerable groups [7]. The OECD has identified young people, women, middle-aged and older individuals, and migrants as vulnerable groups in the labor market [8]. Those workers are also reported to be more likely to leave the labor force owing to unemployment, disability, or economic inactivity [9]. By contrast, civil servants, teachers, and employees of non-profit organizations in Japan have been found to have lower turnover rates and greater job security [10].

It is assumed that individual socioeconomic status is still significantly associated with unemployment during COVID-19. A cross-sectional study conducted in the United States examined adverse outcomes associated with COVID-19 and the country’s stay-home policies. It found that African Americans, Hispanics, women, and low-income households were more likely to experience unemployment, food insecurity, mental health problems, poor access to health care, and rent or mortgage delinquency [11]. However, the relationship between unemployment and socioeconomic status among Japanese during COVID-19 is unclear. The present investigation was a cohort study of the relationship between socioeconomic status and unemployment during COVID-19 among workers in Japan.

## Methods

This prospective cohort study about COVID-19 among Japanese workers was conducted under the Collaborative Online Research on the Novel-coronavirus and Work (CORoNaWork) Project. Details of the study protocol are described elsewhere [12]. Briefly, we administered a baseline questionnaire on December 22–25, 2020 and a follow-up questionnaire on February 18–19, 2021, when Japan was in its third pandemic wave.

For the baseline survey, we recruited 33,087 workers throughout Japan from 605,381 randomly selected panelists registered with an Internet survey company. The inclusion criteria for participants were being currently employed and aged 20–65 years. This study excluded health-care professionals and caregivers. We applied cluster sampling with stratification by sex, job type, and region. We excluded 6051 invalid responses owing to the following: response time <6 minutes; body weight <30 kg; height <140 cm; inconsistent answers to similar questions; and incorrect answers to questions intended to identify fraudulent responses. We distributed the follow-up questionnaire to the 27,036 people with valid responses to the baseline questionnaire. In total, 19,941 participants completed both questionnaires (follow-up rate, 73.8%).

This study was approved by the Ethics Committee of the University of Occupational and Environmental Health, Japan (reference nos. R2-079 and R3-006). Informed consent was obtained from all participants.

### Baseline characteristics

We retrieved the following data from the baseline survey for inclusion as explanatory variables: age; sex; marital status; socioeconomic status (based on annual household income and education); occupation; and job type. We categorized age into the following five groups: 20–29; 30–39; 40–49; 50–59; and ≥60 years. Marital status was classified into four groups: married (working spouse); married (spouse not working); divorced or widowed; and never married. Annual household income was classified into the following six groups: under 2 million; 2–4 million; 4–6 million; 6–8 million; 8–10 million; and >10 million yen. We categorized education into five groups: up to junior high school; up to high school; up to junior college or technical school; up to university; and graduate school. We categorized occupation into 10 groups: general employee; manager; executive manager; public employee, faculty member, or non-profit organization employee; temporary or contract employee; self-employed; small office/home office; agriculture, forestry, or fishing; professional occupation (e.g., lawyer, tax accountant, medical-related work); and other. Job type was classified into three categories: mainly desk work; work mainly involving interpersonal communication; and mainly manual or physical labor.

### Measurement for unemployment

We ascertained unemployment as follows. First, the baseline survey included only people who were employed at the time of response. In the follow-up survey, in answer to the question “Have you changed your place of work since December 2020?” respondents were asked to select one of the following six options: “no change”; “I was transferred to another company”; “I resigned and got a new job right away”; “I stopped working and was unemployed for a while but am now working”; “I stopped working and started a business (e.g., managing a company, running a sole proprietorship, or engaging in self-employment)”; and “I stopped working and am not currently working (including job seeking).” We defined unemployment as participants choosing one of the following: “I resigned and got a new job right away”; “I stopped working and was unemployed for a while but am now working”; “I stopped working and started a business (e.g., managing a company, running a sole proprietorship, or engaging in self-employment)”; or “I stopped working and am not currently working (including job seeking).”

### Statistical analysis

We determined the odds ratios (ORs) of unemployment for socioeconomic factors using a multilevel logistic model for the prefecture of residence. The multivariate model was adjusted for sex and age. We undertook a trend test by conducting the analysis using age, annual household income, and education as continuous variables. We also used the incidence rate of COVID-19 by prefecture as a prefecture-level variable. The multilevel analysis was performed within this incidence rate of COVID-19. We considered *P* values under 0.05 statistically significant. All analyses were conducted using Stata (Stata Statistical Software release SE16.1; StataCorp LLC, College Station, TX, USA).

## Results

The basic characteristics of the respondents appear in Table 1. There were 11,170 men in the sample, accounting for 56% of the total. The mean age was 48.0 years.

**Table 1.** Basic characteristics of the subjects

|  | N (%) |
| --- | --- |
| Number of subjects | 19941 |
| Sex, Men | 11170 (56.0%) |
| Age |  |
| 20-29 | 1055 (5.3%) |
| 30-39 | 3218 (16.1%) |
| 40-49 | 5929 (29.7%) |
| 50-59 | 7095 (35.6%) |
| 60- | 2644 (13.3%) |
| Marital status |  |
| married (spouse is working) | 8125 (40.7%) |
| married (spouse is not working) | 3060 (15.3%) |
| bereaved/divorced | 2036 (10.2%) |
| unmarried | 6720 (33.7%) |
| Annually household income (million JPY) |  |
| <2 | 1288 (6.5%) |
| ≥2 and <4 | 3989 (20.0%) |
| ≥4 and <6 | 4772 (23.9%) |
| ≥6 and <8 | 3967 (19.9%) |
| ≥8 and <10 | 2629 (13.2%) |
| ≥10 | 3296 (16.5%) |
| Education |  |
| Junior high or high school | 5360 (26.9%) |
| Vocational school, junior college or technical school | 4585 (23.0%) |
| University | 8880 (44.5%) |
| Graduate School | 1116 (5.6%) |
| Occupation |  |
| general employee | 9098 (45.6%) |
| manager | 2088 (10.5%) |
| executive manager | 691 (3.5%) |
| public employee, faculty member, or non-profit organization employee | 2010 (10.1%) |
| temporary or contract employee | 2109 (10.6%) |
| self-employed | 1737 (8.7%) |
| small office home office | 304 (1.5%) |
| agriculture, forestry, or fishing | 155 (0.8%) |
| professional occupation (lawyer, tax accountant, medical-related, etc.) | 1246 (6.2%) |
| others | 503 (2.5%) |
| Jobtype |  |
| mainly desk work | 10268 (51.5%) |
| jobs mainly involving interpersonal communication | 4937 (24.8%) |
| mainly manual or physical labor | 4736 (23.8%) |

Table 2 shows the associations of socioeconomic factors with unemployment: 725 (3.6%) workers had experienced unemployment.

**Table 2.**
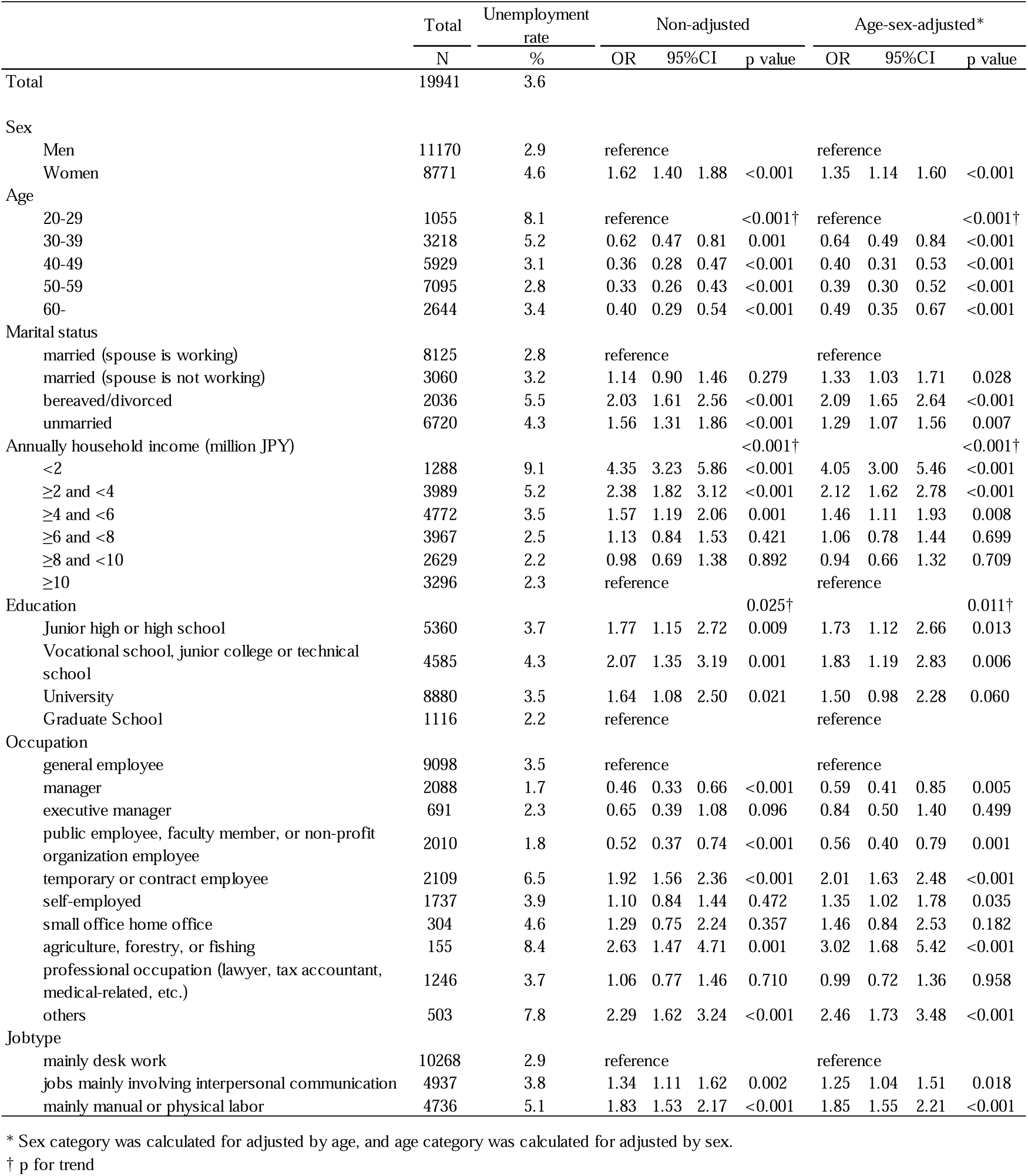
The association between socioeconomic factors and unemployment.

Multivariate analysis showed that the OR of unemployment associated with sex was 1.35 (95% confidence interval [CI], 1.14–1.60) for women compared with men. With increasing age, the OR for unemployment was lower: OR, 0.98; 95% CI, 0.97–0.99; *P* <0.001, adjusted for sex. The respective OR and 95% CI figures for the association with marital status were as follows: 1.33 (1.03–1.71) for being married (spouse not working); 2.09 (1.65–2.64) for being bereaved or divorced; and 1.29 (1.07–1.56) for being unmarried, compared with being married (spouse working). The respective figures for the association with annual household income were as follows: 4.05 (3.00–5.46) for <2 million yen; 2.12 (1.62–2.78) for 2–4 million yen; and 1.46 (1.11–1.93) for 4–6 million yen, compared with >10 million yen. The respective figures for the association with education were as follows: 1.73 (1.12–2.66) for junior high or high school; and 1.83 (1.19–2.83) for vocational school, junior college, or technical school. The respective figures for the association with occupation were as follows: 2.01 (1.63–2.48) for temporary or contract employees; 1.35 (1.02–1.78) for being self-employed; and 3.02 (1.68–5.42) for agriculture, forestry, or fishing, compared with general employees; the figures were 0.56 (0.40–0.79) for public employees, faculty members, or non-profit organization employees. The respective figures for the association with job type were 1.25 (1.04–1.51) for jobs mainly involving interpersonal communication and 1.85 (1.55–2.21) for mainly manual or physical labor, compared with mainly desk work.

## Discussion

Through a prospective cohort study, this investigation examined the association between socioeconomic factors and subsequent unemployment during COVID-19. Under the pandemic of COVID-19, we found that unemployment was associated with socioeconomic factors such as age, sex, marriage, income, education and occupation.

We observed that the risk of unemployment was highest among young people. International Labour Organization (ILO) has reported that worldwide, young people were subjected to the greatest loss of labor opportunities through COVID-19: in 2020, young workers suffered 8.7% job losses compared with 3.7% for adults [13]. The present study conducted in Japan similarly found that young people were more than twice as likely to be unemployed as middle-aged and older workers. This could have been due to the fact that the accommodation and service industry as well as the food and beverage industry (which were most affected by COVID-19) had more young casual workers than other industries [5,14]. In addition, even before the pandemic, there was a high turnover rate of young people in Japan. The turnover rate within 3 years of graduating from university and entering the workforce was around 30% in 2021 [15]. This is thought to be due to their transition from school to work and still looking for the right job.

By industry and occupation, we found the unemployment rate to be higher among temporary and contract workers, agricultural and forestry workers, and manual laborers. Japan’s Labour Force Survey reported that the number of non-regular workers in the country fell sharply under COVID-19 [5]. Many temporary and contract workers were employed in lifestyle-related industries, travel and entertainment services, which were heavily affected by the pandemic [16,17]. Temporary and contract workers are thought to be used to adjust employment and our results clearly confirm this.

We observed a high unemployment rate in the agriculture, forestry, and fisheries sector. The reasons for the high unemployment rate in agriculture are unclear: agriculture is an employment sector that is less susceptible than others to economic fluctuations. One possibility is that the economic downturn in the food service and food industry caused by COVID-19 had an impact on agricultural production [16]. Another explanation for the finding in agriculture, forestry, and fisheries could that it was reflecting a trend for casual work in Japan: temporary employment during the farming season is common, and over 80% of temporary workers have short-term contracts of under one month [18,19]. A further factor could be that the number of farmers in Japan has been declining annually since before COVID-19, and the high unemployment we found in agriculture may not have been due solely to the pandemic [20].

We found that women were more likely to be unemployed than men: the OECD has stated that women are also more vulnerable in society [8]. Japan’s Labour Force Survey during COVID-19 observed a gender difference in the decline in the number of people in employment: women were more likely to be unemployed [5]. The 2021 survey reported that 68% of people in informal employment were women; many of them worked in the accommodation, catering, and lifestyle-related service industries [5]. In the present study, we found that 14.7% of women (compared with 7.4% of men) were in informal employment.

The unemployment rate was also significantly higher for divorced or bereaved people than with dual-earner households. In economic terms, marriage is held to be a rational behavior that seeks economic gain [21,22]. It has long been pointed out that low-income earners and those with unstable employment are less likely to get married [23,24]. The unmarried participants with their precarious employment situation may became unemployed owing to the pandemic. Also, if the divorced or bereaved had a child, it is possible that they were forced to leave the workplace due to school or after-school care leave, because school closures by the pandemic [25].

With regard to income and education, we observed that the lower the income and lower the education, the greater was the likelihood of unemployment. It is widely known that such socially vulnerable groups are at higher risk of unemployment [7,9,26]; we found a similar trend during COVID-19. The impact of unemployment on the lives of those with lower incomes is accordingly greater, and they constitute the group in highest need of social support.

Overall, COVID-19 appears to have increased difficulties for a previously vulnerable group. Previous studies have shown that people of lower socioeconomic status are more likely to face difficulties in the event of a pandemic. However, much of the research has focused on the higher risk of contracting infectious diseases and, as a result, being more likely to face problems such as healthcare costs and unemployment [7,27-30]. Our findings suggest that in the event of a major epidemic, resulting in unemployment among vulnerable segments of the labour market, regardless of whether workers themselves are infected. Thus, there is a need for employment and financial support for socially vulnerable groups in the event of a major epidemic.

There are several limitations of this study. First, it was unclear why the participants had experienced unemployment: we did not know whether it was due to the effects of COVID-19, company bankruptcy or financial difficulties, or the participants’ voluntary decision to change jobs. Second, this study was conducted as an Internet survey; thus, the generalizability of our results is unclear. It is possible that individuals who were genuinely penurious did not have Internet access and could not participate in the survey. If such people had taken part in the survey, the bias would have been stronger. We attempted to reduce subject bias as much as possible by sampling by region and occupation based on infection rates. Third, of the 27,036 individuals who participated in the baseline survey, 7095 did not respond to the follow-up survey (non-participation rate, 26%). That may have led to further bias.

## Conclusion

We confirmed the relationship between socioeconomic factors and subsequent unemployment in the case of COVID-19. There is a need for widespread, sustained support for socially vulnerable groups in the form of both short-term and long-term vocational training and health care.

## Data Availability

Data are available from the corresponding author, Tomohisa Nagata, on request.

## Acknowledgements

The current members of the CORoNaWork Project, in alphabetical order, are as follows: Dr. Yoshihisa Fujino (present chairperson of the study group), Dr. Akira Ogami, Dr. Arisa Harada, Dr. Ayako Hino, Dr. Hajime Ando, Dr. Hisashi Eguchi, Dr. Kazunori Ikegami, Dr. Kei Tokutsu, Dr. Keiji Muramatsu, Dr. Koji Mori, Dr. Kosuke Mafune, Dr. Kyoko Kitagawa, Dr. Masako Nagata, Dr. Mayumi Tsuji, Ms. Ning Liu, Dr. Rie Tanaka, Dr. Ryutaro Matsugaki, Dr. Seiichiro Tateishi, Dr. Shinya Matsuda, Dr. Tomohiro Ishimaru, and Dr. Tomohisa Nagata. All members are affiliated with the University of Occupational and Environmental Health, Japan.

## Contributors

MK wrote the manuscript. TN and AO created the questionnaire, analyzed the data, and reviewed the manuscript, analyzed the data, and provided advice on interpretation. AH, ST, MT, SM, KM reviewed the manuscript. YF reviewed the manuscript and contributed to the overall survey planning, questionnaire creation, and securing funding for research.

## Funding

This word was supported by the research grant from the University of Occupational and Environmental Health, Japan (no grant number); Japanese Ministry of Health, Labour and Welfare (H30-josei-ippan-002, H30-roudou-ippan-007, 19JA1004, 20JA1006, and 210301-1); Anshin Zaidan (no grant number), the Collabo-Health Study Group (no grant number), and Hitachi Systems, Ltd. (no grant number) and scholarship donations from Chugai Pharmaceutical Co., Ltd. (no grant number). The funder was not involved in the study design, collection, analysis, interpretation of data, the writing of this article or the decision to submit it for publication. All authors declare no other competing interests.

## Competing interests

The authors declare no conflicts of interest associated with this manuscript.

## Patient consent for publication

Informed consent was obtained in the form of the website.

## Ethics approval

This study was approved by the ethics committee of the University of Occupational and Environmental Health, Japan (reference No. R2-079 and R3-006).

## Data availability statement

Data are available from the corresponding author, Tomohisa Nagata, on request.

## Notes

### Competing Interest Statement

The authors have declared no competing interest.

### Clinical Trial

NA

### Funding Statement

This study was supported and partly funded by the following: the University of Occupational and Environmental Health, Japan, General Incorporated Foundation (Anshin Zaidan), the Development of Educational Materials on Mental Health Measures for Managers at Small-sized Enterprises, Health, Labour and Welfare Sciences Research Grants, Comprehensive Research for Womens Healthcare (H30-josei-ippan-002), Research for the Establishment of an Occupational Health System in Times of Disaster (H30-roudou-ippan-007), Research for AIDS Policy Research (JPMP20HB1004), scholarship donations from Chugai Pharmaceutical Co., Ltd., the Collabo-Health Study Group, and Hitachi Systems, Ltd.

